# Radiolabeling Molecular Biomarkers of Invasive Pituitary Neuroendocrine Tumors: A Systematic Review

**DOI:** 10.1101/2025.03.21.25324422

**Authors:** Charbel Marche, Julia C. M. Knop, Shivashankar Khanapur, Eric Suero Molina, Mattew McCord, Vanessa Smith, Michael P. Catalino

## Abstract

**Objective:** Pituitary neuroendocrine tumors (PitNETs) are common intracranial neoplasms that can exhibit invasive behavior, leading to increased morbidity, recurrence, and resistance to treatment. Identifying biomarkers associated with tumor invasiveness could improve early diagnosis and guide therapeutic interventions. This systematic review evaluates molecular biomarkers linked to PitNET invasiveness and explores the potential of radiolabeling for noninvasive detection.

**Methods:** A systematic search of PubMed and Embase databases was conducted to identify studies evaluating molecular markers associated with invasive PitNETs. Biomarkers were selected based on their proposed role in tumor invasion, and evidence supporting their clinical relevance was summarized. Additionally, existing radiolabeling techniques for biomarker detection were reviewed.

**Results:** Five key biomarker groups were identified: matrix metalloproteinases (MMPs), urokinase plasminogen activator (uPA) system, myosin 5A (MYO5A), vascular endothelial growth factor (VEGF), and survivin. MMPs were strongly linked to extracellular matrix degradation and invasion, while uPA facilitated invasion via MMP activation. MYO5A and survivin were implicated in epithelial-mesenchymal transition and tumor motility, and VEGF promoted angiogenesis.

Radiolabeling techniques for MMPs, uPA/uPAR, VEGF, and survivin demonstrated feasibility for imaging tumor invasiveness, though limitations such as non-specific tracer accumulation remain.

**Conclusions:** This review highlights the potential of molecular biomarkers in predicting PitNET invasiveness and the emerging role of radiolabeled probes in noninvasive imaging. Future research should focus on validating these biomarkers in longitudinal studies and refining radiolabeling techniques to improve diagnostic accuracy and therapeutic targeting of invasive PitNETs.

## Introduction

### Background

Pituitary adenomas, or pituitary neuroendocrine tumors (PitNETs), are clonal proliferations of specific cell lineages originating in the anterior pituitary gland, as outlined in the WHO Classification of Endocrine and Neuroendocrine Tumors, 5th edition.^1^ PitNETs are one of the most common intracranial neoplasms, with an estimated prevalence of approximately 17% in the general population.^2^ Cross-sectional studies from various countries have reported prevalence rates ranging from 78 to 116 cases per 100,000 individuals, translating to roughly 1 case per 1,000 people.^3,4^ The incidence is higher in women during early adulthood, with a mean age of 48, while men exhibit a significantly increased incidence later in life, with a mean age of 57.^5,6^ Furthermore, across all age groups, men tend to present with larger median tumor sizes.^6^ These tumors are typically benign but often exhibit local invasiveness, leading to incomplete resection, early recurrence, lack of biochemical remission, visual impairment, and complications such as pituitary hormone dysfunction.^7^ Aggressive PitNETs, while rare, are by definition invasive tumors that demonstrate resistance to both first-and second-line therapeutic interventions.^8^ Certain subtypes have been identified as carrying a higher risk of aggressive behavior. These high-risk subtypes include silent corticotroph tumors, sparsely granulated somatotroph tumors, Crooke cell tumors, plurihormonal PIT-1 positive tumors, and lactotroph tumors in male patients.^9^

### Recurrence, morbidity, and mortality

Recurrence rates for pituitary tumors demonstrate significant variability depending on the treatment approach. For example, non-secreting PitNETs managed with surgical resection alone, reported 5-year recurrence rates ranging from 15% to 55%, while 10-year recurrence rates range from 44% and 78%. The addition of adjuvant radiotherapy has consistently been shown in the literature to significantly reduce recurrence rates, highlighting its efficacy in long-term disease control.^10^

Morbidity and mortality in PitNETs are influenced by tumor type, hormonal activity, and treatment outcomes. Non-functioning tumors primarily cause mass effects, with post-treatment recurrence rates up to 78% at 10 years without radiotherapy. Historically, they have been associated with increased sick leave and disability retirement when compared to the general population.^11^ Functioning tumors are associated with increased morbidity and mortality, often due to cardiovascular or metabolic complications. Aggressive subtypes exhibit poor outcomes despite multimodal treatment, underscoring the importance of early diagnosis and tailored therapy to mitigate disease burden and treatment-related sequelae.^8,12^

### Invasion, aggression, and malignancy

Invasion of surrounding structures occurs in 35% of PitNETs, despite that these tumors are considered histologically benign.^13^ This invasion can occur superiorly through the diaphragma sellae, inferiorly into the basal dura towards the sphenoid sinus, or laterally into the cavernous sinuses.^14^ Figure 1 shows a progression of ACTH tumors from early to late diagnosis. Determining whether a particular tumor will become invasive or distinguishing it from a malignant pituitary tumor can be challenging. Metastatic PitNETs, which account for only 0.13% of pituitary tumors, are associated with a markedly higher mortality rate.^15,16^

**Figure 1:**
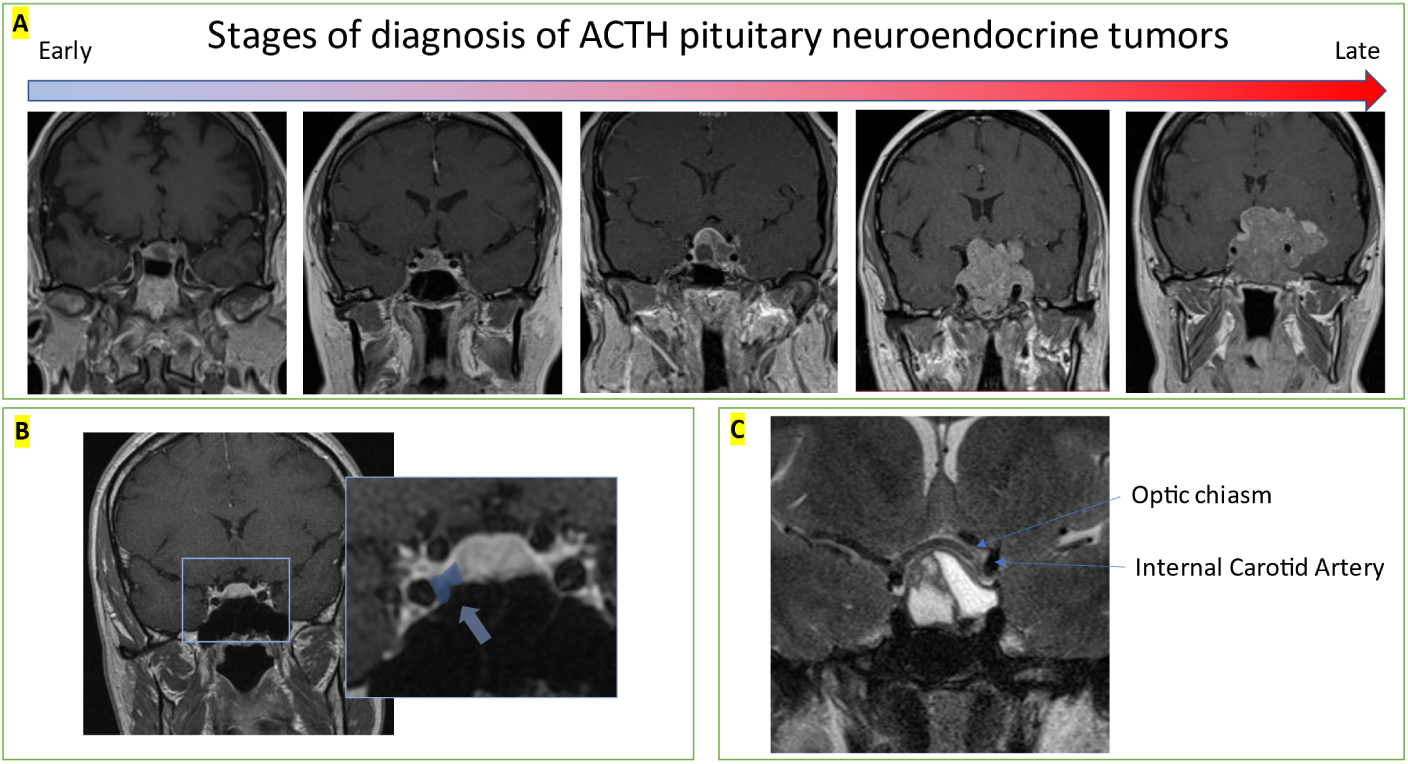
Stages of diagnosis. Panel A shows spectrum of early to late diagnosis of silent ACTII (corticotroph) pituitary neuroendocrine tumors. Left showing early diagnosis of invasion and right showing later diagnosis of highly invasive tumor with, optic nerve compression, vision loss, and brain invasion. Panel H shows an incidentally found right sided lesion with early cavernous sinus Invasion. Panel C shows high resolution T2 Mill which is able to identity critical parasellar structures including optic chiasm and internal carotid artery.

### Radiographic staging/classification

High rates of incidentally found PitNET, as described above, necessitate radiographic classification of invasiveness to predict long-term clinical outcomes as well as biological aggression.

A few commonly used imaging-based grading scales for pituitary tumors are the Knosp, Hardy, and Trouillas classifications. The Knosp classification system consists of grades 0-4 (including 3a and 3b), with higher grades, indicating a greater likelihood of cavernous sinus involvement and lower rates of gross total resection.^17^ The scale has shown strong overall interrater reliability, but this reliability is weaker for middle grades.^18^ Dichotomizing the scale into low (0-2) and high (3-4) grades or further into high (3b-4), medium (2-3a), and low (0-1) improves reliability and clinical utility.^18,19^ However, intermediate grades 2 and 3a continue to present an uncertain prognosis with regard to invasion, as approximately 30-44% of tumors in these categories are found to infiltrate the cavernous sinus.^19^ Studies additionally suggest that the Knosp scale alone may not fully predict tumor behavior and resectability.^20^ A visualization of this grading system in Figure 2 Panel A.^21^ The grading is assigned based on coronal images and the lateral spread of the tumor based on tangents drawn from the carotid arteries as well as the degree of encapsulation of the carotid arteries by the tumor.^21^

**Figure 2:**
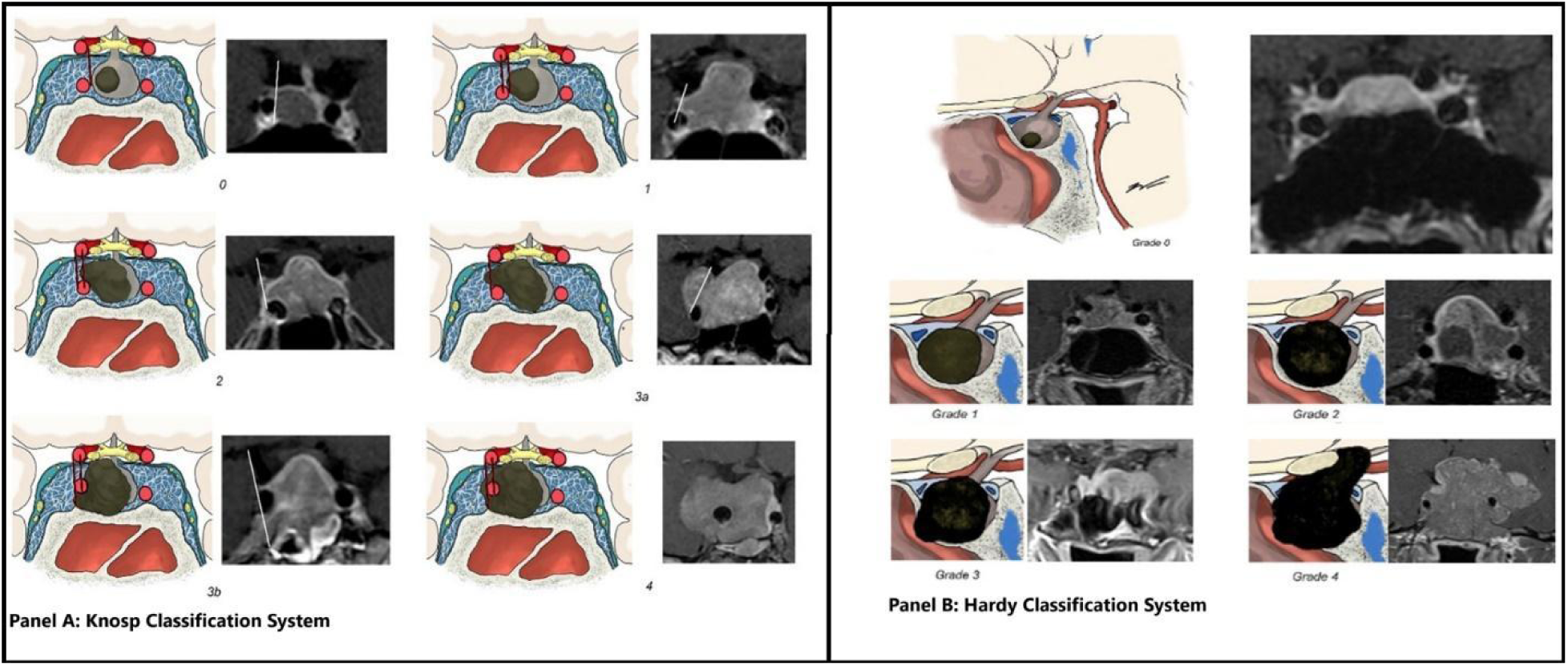
A visualization of the stages of the Knosp and Hardy classifications and accompanying MRI images.

The Hardy-Wilson classification system evaluates the tumor based on the extent of sellar invasion and suprasellar extension, categorizing tumors into grades 1 through 4.^21^ It has been shown to be outperformed by the Knosp scale in predicting surgical outcomes.^21^ A visualization of the Hardy-Wilson grading scale is outlined in Figure 2 Panel B.^21^ The classification is based on the invasion of PitNETs into the sella turcica and into adjacent structures.^21^

The Trouillas classification system leverages both radiological characteristics as well as pathological characteristics to classify PitNETs into one of 5 separate classifications. Grade 1a is reserved for non-invasive, non-proliferative tumors, while grade 1b is defined as non-invasive yet proliferative tumors. Grade 2a tumors are invasive, but not proliferative, while 2b tumors are both invasive and proliferative. Grade 3 tumors are metastatic tumors in which there are cerebrospinal or systemic metastasis. Invasion is determined by histological or radiologic evidence of cavernous or sphenoid sinus invasion. Proliferation is defined as two out of three of the following criteria: Ki-67 >1%, mitoses with n > 2 in 10x high-power field (HPF), or P53 positive in > 10 nuclei in 10x HPF.^22^

Given the high prevalence and rate of invasion/recurrence in young patients, early diagnosis and appropriate management of invasive pituitary neuroendocrine tumors is crucial to prevent long-term morbidity. The problem is that there are no accurate diagnostic methods for determining future malignancy risk. All aggressive PitNETs are invasive by definition so appropriately quantifying invasiveness could help develop appropriate long-term prognostic biomarkers.

Although existing classification systems offer valuable guidance for managing pituitary tumors, they do not predict whether a tumor has invasive/malignant *potential*; they only assess its current invasive status. Timely intervention is critical. Studies have shown that analysis of biomarkers in the post-operative pathologic setting can predict post-operative complete remission or tumor progression.^22^ Consequently, this paper aims to review the current literature and explore potential radiologic biomarkers that may help better predict which patients could benefit from more aggressive early treatment.

## Methods

We performed a comprehensive literature review focusing on five groupings of biomarkers potentially linked to local tissue invasion seen in PitNETs.

Relevant papers were identified through a systematic search of the Embase and PubMed databases. The searches, conducted in October 2024, targeted studies with the following terms listed in Table 1.

For each biomarker, we summarize the evidence supporting its association with invasiveness and discuss the proposed mechanisms underlying its role in tumor progression. The cited <u>database searching tool</u> was used to search over all available publication years and retrieve unique papers.^23^ Studies were then screened for inclusion based on their analysis of the correlation or association between the biomarker expression and PitNETs invasiveness. Meta-analyses were prioritized for review, and any individual studies included within these meta-analyses were excluded from further analysis to prevent redundancy. The evaluation of the availability and applicability of radiolabeling techniques for their detection concludes this review.

## Results

### Matrix Metalloproteinases

#### MMP promotes tissue invasion through the degeneration of type IV collagen and promotes angiogenesis

Matrix metalloproteinases (MMPs) are thought to promote pituitary tumor invasion via multiple mechanisms. The primary proposed mechanism involves the degradation of the extracellular matrix (ECM), compromising its structural integrity, and thus allowing for invasion.^24^ The dura mater—largely composed of type IV collagen—is particularly susceptible, as MMP-9 is a type IV collagenase.^24^ Additionally, multiple MMPs have been positively associated with angiogenesis, further contributing to tumor progression.^25^

Given the established association between elevated MMP expression and invasive PitNETs, one study hypothesized that this increased expression could be linked to heightened DNA methylation of promotors of tissue inhibitors of matrix metalloproteinases (TIMPs), a phenomenon they also observed in invasive pituitary tumors.^26^ This is demonstrated in Figure 3.^27^ The combination of relatively elevated MMP levels as well as decreased TIMP levels is proposed to lead to increased degradation of collagen of the ECM, allowing for tumor invasion.

**Figure 3:**
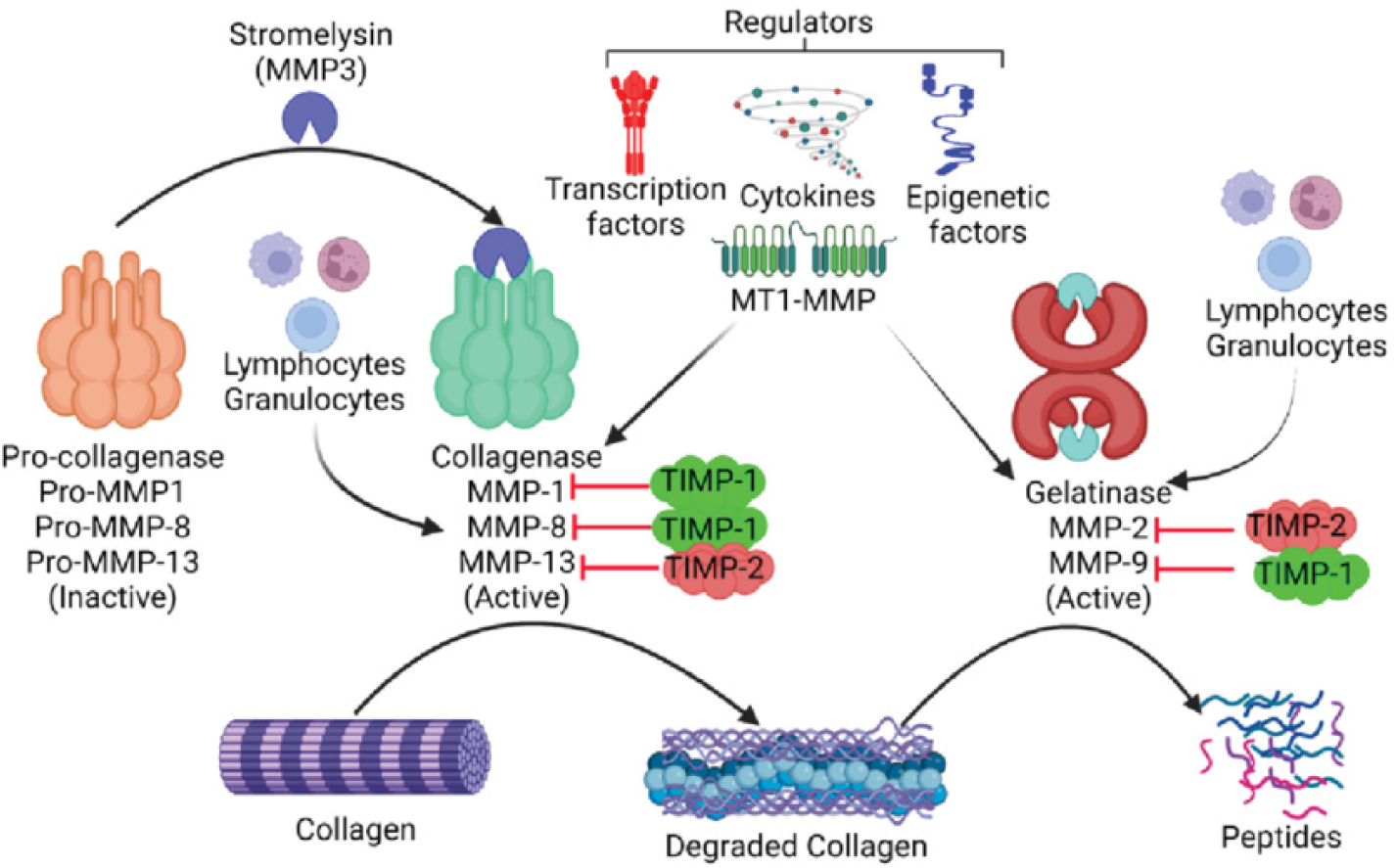
Regulation of collagen degradation by MMPs.

Metalloproteinases (MMPs) are some of the more well-studied potential markers for tumor invasion. Multiple studies have demonstrated significantly higher expression of MMP-9 and/or MMP-2, among others, in invasive PitNETs compared to non-invasive tumors. The results of a literature review are described in detail in Table 2. All studies showed a statistically significant association between MMP-9 and invasiveness, as determined by multiple methods. The included studies covered 2,699 cases of pituitary tumors. MMP-2 was discussed in a single meta-analysis which also found that there was a significant association between MMP-2 presence in tumor and invasiveness.^25,28–31^ In conclusion, MMPs are clearly activated in pituitary neuroendocrine tumor invasion, however, the exact mechanism remains unclear.

#### Summary of radiolabels for MMP

In a 2001 study, researchers outlined the methodology for synthesizing the radiolabeled compound ^111^In−DTPA−N-TIMP-2, designed to bind and inhibit MMP-3. This radiolabel was created by conjugating the MMP inhibitor TIMP-2 with diethylenetriamine pentaacetic acid (DTPA) and then radiolabeling it with indium-111. The resulting compound demonstrated serum stability over 48 hours, with approximately 5% of the radiolabel dissociating during this time. Furthermore, the radiolabel retained its inhibitory activity, was pyrogen-free, and met sterility standards.^32^ Given the established association between pituitary tumors and elevated levels of MMP-9—and, to a lesser extent, MMP-2—there is potential for further research to develop radiolabels targeting these specific molecules.

A 2014 study reported the development of activatable cell-penetrating probes designed to assess MMP-2 and MMP-9 activity, though in the distinct context of post-myocardial infarction cardiac remodeling in murine models. These probes were engineered to be cell-permeable and activatable upon interaction with MMPs, enabling real-time imaging of enzymatic activity.^33^ While not applied to PitNETs, the methodology highlights a potential avenue for utilizing similar probes to evaluate MMP activity in the context of invasive pituitary tumors, offering new insights into tumor behavior and potential diagnostic strategies.

### Urokinase-Type Plasminogen Activator (uPA)

#### uPAR is an upstream receptor that promotes invasiveness through activation of MMPs

The uPA system is proposed to contribute to multiple processes involved in carcinogenesis, tumor invasion, and metastasis. Its primary function is the conversion of plasminogen to plasmin, which facilitates the proteolytic degradation of the basement membrane and extracellular matrix through the activation of MMPs, thus allowing invasion and metastasis.^34^ Moreover, the uPA system has been shown to promote epithelial-mesenchymal transition, activate mitogenic signaling pathways, inhibit apoptosis, and enhance cell migration, invasion, and metastasis.^34^ Figure 4 includes a visual representation of the uPA system.^35^

**Figure 4:**
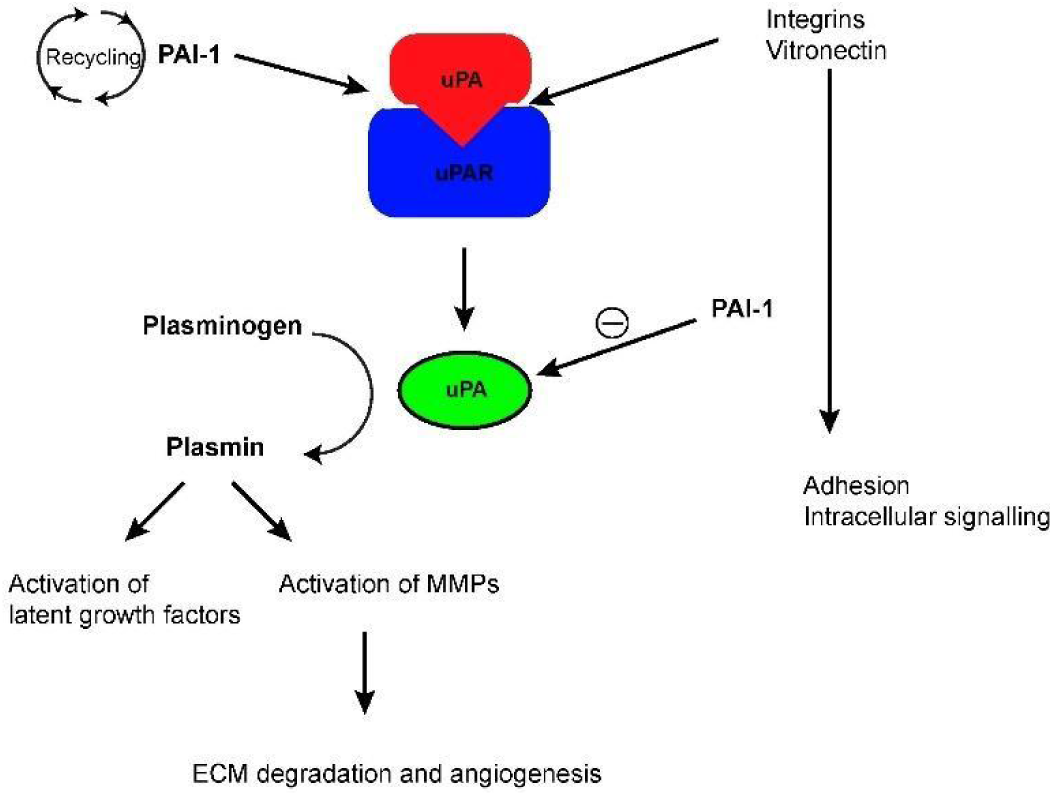
Regulation of uPA and activation of plasmin.

The uPA system has been widely reported in the literature to be associated with tumor invasiveness and metastatic potential across various malignancies, with significant prognostic implications.^36^

#### uPA overexpression is weakly associated with pituitary tumor invasiveness

A 2003 blinded study examined the expression of uPA and uPA receptor (uPAR) concerning the invasiveness of both functional and nonfunctional pituitary tumors. Samples were collected from 84 patients at the University Medical Center Hamburg-Eppendorf during transsphenoidal surgery. The patients had a mean age of 47 years, ranging from 17 to 79 years. Tissue expression of uPA was assessed using immunohistochemical staining, and four independent, blinded observers rated the expression on the following scale: “distinct” for >50% of positively stained cells, “moderate” for <50%, “slightly positive” for small clusters or weak diffuse reactions, and “negative.”^37^

uPA and uPAR were expressed in 89% and 90% of pituitary tumors, respectively. Staining was positive in 90% and 92% of invasive tissues for uPA and uPAR, respectively, and in 88% of non-invasive tissues for both markers. Invasiveness was determined via preoperative MRI and verified by intraoperative inspection. Expression levels of uPA and uPAR did not correlate with invasiveness in functional pituitary tumors, though uPA showed a stronger tendency toward overexpression in invasive compared to non-invasive non-functional tumors (*Chi-square P=0.053*). uPAR did not show any association with invasiveness.^37^

#### Summary of radiolabels for uPA and uPAR

Several successful attempts have been made to conjugate DOTA (1,4,7,10-tetraazacyclododecane-1,4,7,10-tetraacetic acid) and NODAGA (1-(1-carboxy-3-carboxy-propyl)-4,7-(carboxy-methyl)-1,4,7-triazacyclononane) with various radionuclides to target uPAR in mouse models.^38^ However, these studies have reported confounding levels of non-specific baseline tracer accumulation in both liver and tumor tissue, particularly with ^64^Cu-labeled markers.^38^

### Myosin 5a (MYO5A)

#### MYO5A expression is upregulated when bound to Snail and Akt and associated with invasion through the epithelial-mesenchymal transition (EMT) pathway

MYO5A is an unconventional myosin protein that is upregulated by EMT markers such as Snail and Akt2. It is additionally proposed that Snail could be a transcriptional activator of MYO5A. EMTs promote the loss of epithelial characteristics of epithelial cells and induce the gain of mesenchymal characteristics which allows for loose adherence and increased motility. These EMTs, and MYO5A by extension, are associated with increased invasiveness and metastasis.^39,40^

Increased expression of MYO5A has been observed in several metastatic cell lines, including metastatic colorectal, lung, breast, and prostate cancers, as well as in invasive and metastatic esophageal squamous cell carcinomas.^39,40^

#### Myosin 5a gene expression is elevated in invasive pituitary tumors

A 2010 study analyzed a sample of 40 non-functional PitNETs resected via transsphenoidal surgery at Bicêtre Hospital in France. The study assessed gene expression and protein levels using RT-PCR and immunohistochemical (IHC) staining, respectively, including the MYO5A gene and protein. Tumor invasiveness was classified based on preoperative and intraoperative MRI findings, employing the modified Hardy grading scale. Grade III and IV tumors were categorized as invasive. Of the tumors evaluated, 22 were classified as invasive, while the remaining 18 were not.^41^ The study demonstrated that MYO5A gene expression was significantly elevated in invasive tumors compared to noninvasive tumors, with a corresponding increase observed at the protein level.^41^

#### Summary of Radiolabels

As of the time of this paper’s writing, research on radiolabels targeting MYO5A remains limited.

### Vascular Endothelial Growth Factor (VEGF)

#### Hypoxia-induced VEGF upregulation promotes angiogenesis

Tumor hypoxia is thought to upregulate VEGF expression, promoting its release within the tumor microenvironment.^42^ VEGF and its receptor are known to trigger angiogenesis and are correlated with invasiveness and metastaticity, but the mechanisms by which VEGF/VEGFR1 may promote these tumor characteristics remain unknown.^43^

VEGF and VEGF receptor 1 (VEGFR1) expression has been documented to be associated with invasion and metastasis for various types of cancers including head and neck squamous cell cancer, and bladder cancer, among others.^42,44,45^

Nine studies are outlined in Table 3 below with their respective results. Six of these nine studies demonstrated positive results for increased VEGF expression in invasive pituitary tumors, while the remaining 3 were not able to identify any statistically significant difference between the two groups. These studies included 450 patients in total.^46–54^ The variability in findings regarding the association of VEGF with invasion suggests that it may not be the most sensitive biomarker for assessing invasive potential.

#### Radiolabels for Vascular Endothelial Growth Factor and Receptor

Multiple studies have explored the development of radiolabels targeting VEGFR1. These include ^99^mTc-labeled VEGF165 for SPECT imaging,^55^ 177Lu-labeled anti-VEGFR1 antibodies for radioimmunotherapy^56^, and ^89^Zr-labeled single-chain VEGF mutants for selective PET imaging of VEGFR-1 and VEGFR-2^57^. ^64^Cu-labeled VEGF121 has demonstrated utility for in vivo visualization of VEGFR expression^58^, while ^68^Ga-labeled peptides and other small molecule VEGFR inhibitors targeting VEGFRs show promise for PET imaging.^59,60^ Radiolabeled bevacizumab, an anti-VEGF antibody, has additionally been developed for PET/SPECT imaging of VEGF in tumors using ^89^Zr and 111In.^61^

### Survivin

#### Survivin promotes tumor invasion by preventing cell death and promoting cell motility

Survivin is thought to inhibit cell apoptosis and thereby contribute to tumor cell proliferation. Its role in promoting invasion is believed to involve multiple mechanisms. Research has shown that survivin enhances oxidative phosphorylation and mitochondrial repositioning, facilitating cellular respiration to generate the energy required for tumor cell migration and invasion.^62^ Furthermore, another study demonstrated that the formation of a survivin-XIAP complex within tumor tissue promotes cell motility, contributing to invasion and metastasis.^63^

Increased expression of survivin has been found to be increased in a variety of malignant tumors including lung cancer, breast cancer, and gliomas.^64^

#### Survivin expression is associated with pituitary tumor invasion

A 2017 meta-analysis examined 9 studies conducted across Asia and Europe, involving a total of 489 patients, 266 of whom had invasive pituitary tumors. Survivin expression was associated with tumor invasiveness in these studies, with 8 using IHC staining and 1 using RT-PCR. Positive survivin expression was defined as staining in the nucleus or cytoplasm. Of the 9 studies, 7 showed statistically significant associations between survivin expression and increased invasiveness. This resulted in an odds ratio of 6.23, with a 95% confidence interval of 3.97 to 9.76 (P < 0.001).^64^

#### Radiolabels for survivin

Several radiolabeled compounds targeting survivin have been developed for tumor imaging, including radioiodinated 4,6-diaryl-3-cyano-2-pyridinone derivatives^65^ and 3-phenethyl-2-indolinone derivatives^66^. These compounds demonstrated specific binding to survivin in vitro murine models and showed moderate tumor uptake in vivo.^66^

## Discussion

The findings from our review underscore the critical role of matrix metalloproteinases (MMPs) in the invasion of PitNETs. Elevated MMP expression correlates with increased tumor invasiveness, likely through extracellular matrix degradation and angiogenesis promotion. The observation that DNA methylation of TIMP promoters may contribute to unchecked MMP activity further supports the hypothesis that the MMP-TIMP balance plays a key role in tumor progression. Despite the compelling evidence linking MMPs to invasion, the exact regulatory mechanisms remain unclear, warranting further research into their upstream signaling pathways and interactions.

Beyond MMPs, the uPA and uPAR system has been implicated in tumor invasion through its activation of MMPs and facilitation of EMT. However, direct association with pituitary tumor invasiveness remains inconclusive. The single 2003 study observing invasiveness with the presence of these biomarkers determined invasiveness via MRI and surgeon observation, rather than the gold standard of pathologic analysis. Novel research observing uPA or uPAR expression with pathologic invasion may be beneficial and give more conclusive results.

Myosin 5a (MYO5A) emerges as another promising marker, with studies demonstrating its upregulation in invasive pituitary tumors. MYO5A’s interaction with EMT markers such as Snail and Akt further supports its role in facilitating tumor cell motility and metastasis. However, research on MYO5A remains limited highlighting the need for additional studies to validate its potential as a diagnostic or therapeutic target.

Vascular endothelial growth factor (VEGF) has been widely studied for its role in tumor angiogenesis. Although several studies demonstrate an association between VEGF upregulation and pituitary tumor invasion, findings are inconsistent, with some failing to identify a statistically significant correlation. This suggests that VEGF may not be the most reliable independent predictor of invasiveness in pituitary tumors.

Survivin, a well-established inhibitor of apoptosis, appears to contribute to tumor invasion by enhancing cell motility and metabolic adaptation. Meta-analysis findings indicate a strong association between survivin expression and pituitary tumor invasiveness, with significant odds ratios supporting its prognostic value. Given its involvement in multiple malignancies, survivin presents as a compelling target for future research into invasive pituitary tumors.

The exploration of radiolabeled compounds for imaging and therapeutic targeting of these markers represents an exciting frontier in neuro-oncology. Radiolabeled inhibitors for MMPs, uPA/uPAR, VEGF, and survivin have demonstrated feasibility, with several compounds showing promising tumor specificity and in vivo stability. However, challenges such as non-specific tracer accumulation, particularly with 64Cu-labeled uPAR markers, indicate the need for further refinement of radiolabeling strategies.

## Conclusions

In conclusion, this review highlights the multifactorial nature of pituitary tumor invasion, driven by MMPs, the uPA system, MYO5A, VEGF, and survivin. While each marker contributes to tumor progression through distinct mechanisms, their combined effects likely orchestrate the invasive phenotype. Future research should focus on elucidating the interplay between these pathways, developing targeted imaging agents, and refining therapeutic interventions to mitigate tumor invasion and improve patient outcomes.

### Limitations

A major limitation of the research discussed is that these studies overwhelmingly establish the association of already known and observed invasiveness, as defined by their respective methods, with biomarker presence. Whether these biomarkers are elevated in tumors that have not yet become invasive but will be, has not been investigated and would require longitudinal studies to establish sufficient evidence.

Another significant limitation is the method in which the invasiveness is pituitary tumors is determined. Many studies determine invasiveness via non-pathological methods such as imaging and surgeon observations. Both methods of determining invasiveness are subject to error and may affect the accuracy of study results and are not considered the gold standard for determining invasiveness. Thus, associations or lack thereof may be erroneous due to the potentially inaccurate classification of tumors into invasive and non-invasive subgroups.

### Summary of Results and Future Directions

This paper reviews the evidence linking biomarkers—MMP-2, MMP-9, uPA, MYO5A, VEGF/VEGFR, and survivin—with invasive behavior in PitNETs. These markers are proposed as potential risk indicators and may provide insights into the invasive potential of tumors.

Overwhelmingly, published literature has reported associations between these biomarkers and invasion. Emerging evidence also highlights the feasibility of radiolabeling certain biomarkers, suggesting a promising avenue for noninvasive assessment and prediction of tumor invasiveness, particularly in incidentally discovered pituitary lesions. These findings underscore the potential for biomarker-driven strategies in managing pituitary tumors.

The identification and utilization of biomarkers such as matrix metalloproteinases (MMPs), urokinase plasminogen activator and its receptor (uPA/uPAR), myosin 5A (MYO5A), survivin, and vascular endothelial growth factor (VEGF) have the potential to significantly enhance the prediction of pituitary tumor invasiveness. These markers have demonstrated associations with the invasive behavior of pituitary tumors across both functional and non-functional subtypes. Further investigation into the relationship between these biomarkers (and others) and tumor invasiveness is critical to building a robust dataset that could inform the development of targeted radiolabels.

Although many radiolabels targeting these biomarkers are in the early phases of development, they hold promise for refining diagnostic algorithms and improving the management and surveillance of pituitary tumors. The utility of these radiolabels as reliable predictors of future tumor aggressiveness and invasiveness will require validation through clinical studies as they become more widely available. If these radiolabels are found to be accurate and specific predictors of invasiveness, they may also be used for the development of therapeutic interventions targeting abnormal tissue.

During the course of this systematic review, several other biomarkers potentially associated with the invasiveness of PitNETs have been identified. These included MMP-1, MMP-14, PTTG, reticulin, laminins, type IV collagen, EHZ2, estrogen receptor, HIF1, FBGF-2, LAMA2, DDR1, CLIC2, TOPO 2A, and others. While this analysis focuses on five groups of selected biomarkers with therapeutic potential, further exploration of additional biomarkers is strongly recommended to enhance our understanding of tumor behavior.

Moreover, a logical extension of this discussion involves the investigation of biomarkers specific to pituitary metastatic neuroendocrine tumors. Given the rarity of these conditions, the available data is expected to be limited and in need of longitudinal cohort studies, however, identifying reliable biomarkers for the detection and characterization of metastatic PitNET holds significant potential for advancing the diagnosis and management of these aggressive neoplasms.

## Data Availability

All studies available in public research databases

## Acknowledgments

N/A

## Notes

### Competing Interest Statement

The authors have declared no competing interest.

### Clinical Protocols

https://github.com/Pituitary-Normal-Space/database-searcher

### Funding Statement

This study did not receive any funding

## References

1. Asa SL, Mete O, Perry A, Osamura RY. Overview of the 2022 WHO Classification of Pituitary Tumors. Endocr Pathol. 2022;33(1):6-26. doi:10.1007/s12022-022-09703-7

2. Ezzat S, Asa SL, Couldwell WT, et al. The prevalence of pituitary adenomas: a systematic review. Cancer. 2004;101(3):613–619. doi:10.1002/cncr.20412

3. Daly AF, Beckers A. The Epidemiology of Pituitary Adenomas. Endocrinol Metab Clin North Am. 2020;49(3):347–355. doi:10.1016/j.ecl.2020.04.002

4. Karavitaki N. Prevalence and incidence of pituitary adenomas. Ann Endocrinol. 2012;73(2):79–80. doi:10.1016/j.ando.2012.03.039

5. Castellanos LE, Gutierrez C, Smith T, Laws ER, Iorgulescu JB. Epidemiology of common and uncommon adult pituitary tumors in the U.S. according to the 2017 World Health Organization classification. Pituitary. 2022;25(1):201-209. doi:10.1007/s11102-021-01189-6

6. McDowell BD, Wallace RB, Carnahan RM, Chrischilles EA, Lynch CF, Schlechte JA. Demographic Differences in Incidence for Pituitary Adenoma. Pituitary. 2011;14(1):23–30. doi:10.1007/s11102-010-0253-4

7. Lefevre E, Chasseloup F, Hage M, Chanson P, Buchfelder M, Kamenický P. Clinical and therapeutic implications of cavernous sinus invasion in pituitary adenomas. Endocrine. 2024;85(3):1058–1065. doi:10.1007/s12020-024-03877-2

8. Burman P, Casar-Borota O, Perez-Rivas LG, Dekkers OM. Aggressive Pituitary Tumors and Pituitary Carcinomas: From Pathology to Treatment. J Clin Endocrinol Metab. 2023;108(7):1585–1601. doi:10.1210/clinem/dgad098

9. Tritos NA, Miller KK. Diagnosis and Management of Pituitary Adenomas: A Review. JAMA. 2023;329(16):1386–1398. doi:10.1001/jama.2023.5444

10. Losa M, Picozzi P, Motta M, Valle M, Franzin A, Mortini P. The role of radiation therapy in the management of non-functioning pituitary adenomas. J Endocrinol Invest. 2011;34(8):623–629. doi:10.3275/7618

11. Jonsson B, Nilsson B. The impact of pituitary adenoma on morbidity. Increased sick leave and disability retirement in a cross-sectional analysis of Swedish national data. PharmacoEconomics. 2000;18(1):73–81. doi:10.2165/00019053-200018010-00008

12. Ostrom QT, Price M, Neff C, et al. CBTRUS Statistical Report: Primary Brain and Other Central Nervous System Tumors Diagnosed in the United States in 2015–2019. Neuro-Oncol. 2022;24(Supplement_5):v1-v95. doi:10.1093/neuonc/noac202

13. Scheithauer BW, Kovacs KT, Laws ER, Randall RV. Pathology of invasive pituitary tumors with special reference to functional classification. J Neurosurg. 1986;65(6):733–744. doi:10.3171/jns.1986.65.6.0733

14. Buchfelder M, Fahlbusch R, Adams EF, Roth M, Thierauf P. Growth Characteristics and Proliferation Parameters of Invasive Pituitary Adenomas. In: Piscol K, Klinger M, Brock M, eds. Neurosurgical Standards Cerebral Aneurysms Malignant Gliomas. Springer; 1992:381–386. doi:10.1007/978-3-642-77109-5_67

15. Hansen TM, Batra S, Lim M, et al. Invasive adenoma and pituitary carcinoma: a SEER database analysis. Neurosurg Rev. 2014;37(2):279–286. doi:10.1007/s10143-014-0525-y

16. Blevins LS, Verity DK, Allen G. Aggressive pituitary tumors. Oncol Williston Park N. 1998;12(9):1307–1312, 1315; discussion 1315-1318.

17. Micko A, Oberndorfer J, Weninger WJ, et al. Challenging Knosp high-grade pituitary adenomas. J Neurosurg. 2019;132(6):1739–1746. doi:10.3171/2019.3.JNS19367

18. Mooney MA, Hardesty DA, Sheehy JP, et al. Interrater and intrarater reliability of the Knosp scale for pituitary adenoma grading. J Neurosurg. 2017;126(5):1714–1719. doi:10.3171/2016.3.JNS153044

19. Fang Y, Pei Z, Chen H, et al. Diagnostic value of Knosp grade and modified Knosp grade for cavernous sinus invasion in pituitary adenomas: a systematic review and meta-analysis. Pituitary. 2021;24(3):457–464. doi:10.1007/s11102-020-01122-3

20. Enseñat J, Ortega A, Topcewski T, et al. [Predictive value of the Knosp classification in grading the surgical resection of invasive pituitary macroadenomas. A prospective study of 23 cases]. Neurocir Astur Spain. 2006;17(6):519-526.

21. Araujo-Castro M, Acitores Cancela A, Vior C, Pascual-Corrales E, Rodríguez Berrocal V. Radiological Knosp, Revised-Knosp, and Hardy–Wilson Classifications for the Prediction of Surgical Outcomes in the Endoscopic Endonasal Surgery of Pituitary Adenomas: Study of 228 Cases. Front Oncol. 2022;11:807040. doi:10.3389/fonc.2021.807040

22. Trouillas J, Roy P, Sturm N, et al. A new prognostic clinicopathological classification of pituitary adenomas: a multicentric case-control study of 410 patients with 8 years post-operative follow-up. Acta Neuropathol (Berl). 2013;126(1):123–135. doi:10.1007/s00401-013-1084-y

23. Marche C. Literature Review Search Tool. Published online October 2024. Accessed February 21, 2025. https://github.com/Pituitary-Normal-Space/database-searcher

24. Yang C, Bao X, Wang R. Role of matrix Metalloproteinases in pituitary adenoma invasion. Chin Neurosurg J. 2018;4(1):2. doi:10.1186/s41016-017-0109-0

25. Pan L xiong, Chen Z ping, Liu Y sheng, Zhao J hong. Magnetic resonance imaging and biological markers in pituitary adenomas with invasion of the cavernous sinus space. J Neurooncol. 2005;74(1):71–76. doi:10.1007/s11060-004-6150-9

26. Yang Y, Huang F, Wu X, Huang C, Li Y. Demethylation of TIMP2 and TIMP3 Inhibits Cell Proliferation, Migration, and Invasion in Pituitary Adenomas. Discov Med. 2024;36(184):971–980. doi:10.24976/Discov.Med.202436184.90

27. Matrix Metalloproteinases and Their Inhibitors. Accessed November 25, 2024. https://encyclopedia.pub/entry/6088

28. Liu HY, Gu WJ, Wang CZ, Ji XJ, Mu YM. Matrix metalloproteinase-9 and-2 and tissue inhibitor of matrix metalloproteinase-2 in invasive pituitary adenomas: A systematic review and meta-analysis of case–control trials. Medicine (Baltimore). 2016;95(24):e3904. doi:10.1097/MD.0000000000003904

29. Sun B, Dai C, Liu X, Wang R, Kang J. Matrix Metalloproteinase 9 for Evaluating Invasiveness and Recurrence in Pituitary Adenoma Patients: A Meta-analysis. Published online February 19, 2020. doi:10.21203/rs.2.24007/v1

30. Guo H, Sun Z, Wei J, et al. Expressions of Matrix Metalloproteinases-9 and Tissue Inhibitor of Metalloproteinase-1 in Pituitary Adenomas and Their Relationships with Prognosis. Cancer Biother Radiopharm. 2019;34(1):1–6. doi:10.1089/cbr.2018.2589

31. Gong J, Zhao Y, Abdel-Fattah R, et al. Matrix metalloproteinase-9, a potential biological marker in invasive pituitary adenomas. Pituitary. 2008;11(1):37–48. doi:10.1007/s11102-007-0066-2

32. Giersing BK, Rae MT, CarballidoBrea M, Williamson RA, Blower PJ. Synthesis and Characterization of 111In−DTPA−N-TIMP-2: A Radiopharmaceutical for Imaging Matrix Metalloproteinase Expression. Bioconjug Chem. 2001;12(6):964–971. doi:10.1021/bc010028f

33. van Duijnhoven SMJ, Robillard MS, Hermann S, et al. Imaging of MMP activity in postischemic cardiac remodeling using radiolabeled MMP-2/9 activatable peptide probes. Mol Pharm. 2014;11(5):1415–1423. doi:10.1021/mp400569k

34. Mekkawy AH, Pourgholami MH, Morris DL. Involvement of Urokinase-Type Plasminogen Activator System in Cancer: An Overview. Med Res Rev. 2014;34(5):918–956. doi:10.1002/med.21308

35. Leurer C, Rabbani SA, Leurer C, Rabbani SA. Plasminogen Activator System — Diagnostic, Prognostic and Therapeutic Implications in Breast Cancer. In: A Concise Review of Molecular Pathology of Breast Cancer. IntechOpen; 2015. doi:10.5772/59429

36. Reuning U, Magdolen V, Wilhelm O, et al. Multifunctional potential of the plasminogen activation system in tumor invasion and metastasis (review). Int J Oncol. 1998;13(5):893–1799. doi:10.3892/ijo.13.5.893

37. Knappe UJ, Hagel C, Lisboa BW, Wilczak W, Lüdecke DK, Saeger W. Expression of serine proteases and metalloproteinases in human pituitary adenomas and anterior pituitary lobe tissue. Acta Neuropathol (Berl). 2003;106(5):471–478. doi:10.1007/s00401-003-0747-5

38. Ploug M. Structure-Driven Design of Radionuclide Tracers for Non-Invasive Imaging of uPAR and Targeted Radiotherapy. The Tale of a Synthetic Peptide Antagonist. Theranostics. 2013;3(7):467–476. doi:10.7150/thno.3791

39. Sato N, Fujishima F, Nakamura Y, et al. Myosin 5a regulates tumor migration and epithelial-mesenchymal transition in esophageal squamous cell carcinoma: utility as a prognostic factor. Hum Pathol. 2018;80:113–122. doi:10.1016/j.humpath.2018.06.002

40. Lan L, Han H, Zuo H, et al. Upregulation of myosin Va by Snail is involved in cancer cell migration and metastasis. Int J Cancer. 2010;126(1):53–64. doi:10.1002/ijc.24641

41. Galland F, Lacroix L, Saulnier P, et al. Differential gene expression profiles of invasive and non-invasive non-functioning pituitary adenomas based on microarray analysis. Endocr Relat Cancer. 2010;17(2):361–371. doi:10.1677/ERC-10-0018

42. Yang Y, Cao Y. The impact of VEGF on cancer metastasis and systemic disease. Semin Cancer Biol. 2022;86:251–261. doi:10.1016/j.semcancer.2022.03.011

43. Liu W, Xu J, Wang M, Wang Q, Bi Y, Han M. Tumor-derived vascular endothelial growth factor (VEGF)-A facilitates tumor metastasis through the VEGF-VEGFR1 signaling pathway. Int J Oncol. 2011;39(5):1213–1220. doi:10.3892/ijo.2011.1138

44. Kopparapu PK, Boorjian SA, Robinson BD, et al. Expression of VEGF and its receptors VEGFR1/VEGFR2 is associated with invasiveness of bladder cancer. Anticancer Res. 2013;33(6):2381–2390.

45. Sauter ER, Nesbit M, Watson JC, Klein-Szanto A, Litwin S, Herlyn M. Vascular endothelial growth factor is a marker of tumor invasion and metastasis in squamous cell carcinomas of the head and neck. Clin Cancer Res Off J Am Assoc Cancer Res. 1999;5(4):775–782.

46. Tanase C, Codrici E, Popescu ID, et al. Angiogenic markers: molecular targets for personalized medicine in pituitary adenoma. Pers Med. 2013;10(6):539–548. doi:10.2217/pme.13.61

47. Sato M, Tamura R, Tamura H, et al. Analysis of Tumor Angiogenesis and Immune Microenvironment in Non-Functional Pituitary Endocrine Tumors. J Clin Med. 2019;8(5):695. doi:10.3390/jcm8050695

48. Baldys-Waligorska A, Wierzbicka I, Sokolowski G, Adamek D, Golkowski F. Markers of proliferation and invasiveness in somatotropinomas. Endokrynol Pol. 2018;69(2):182–189. doi:10.5603/EP.a2018.0001

49. He W, Huang L, Shen X, et al. Relationship between RSUME and HIF-1α/VEGF-A with invasion of pituitary adenoma. Gene. 2017;603:54–60. doi:10.1016/j.gene.2016.12.012

50. Yilmaz M, Vural E, Koc K, Ceylan S. Cavernous sinus invasion and effect of immunohistochemical features on remission in growth hormone secreting pituitary adenomas. Turk Neurosurg. 2015;25(3):380–388. doi:10.5137/1019-5149.JTN.9347-13.1

51. Sánchez-Ortiga R, Sánchez-Tejada L, Moreno-Perez O, Riesgo P, Niveiro M, Picó Alfonso AM. Over-expression of vascular endothelial growth factor in pituitary adenomas is associated with extrasellar growth and recurrence. Pituitary. 2013;16(3):370–377. doi:10.1007/s11102-012-0434-4

52. Yarman S, Kurtulmus N, Canbolat A, Bayindir C, Bilgic B, Ince N. Expression of Ki-67, p53 and vascular endothelial growth factor (VEGF) concomitantly in growth hormone-secreting pituitary adenomas; which one has a role in tumor behavior ? Neuro Endocrinol Lett. 2010;31(6):823-828.

53. Borg SA, Kerry KE, Royds JA, Battersby RD, Jones TH. Correlation of VEGF production with IL1 alpha and IL6 secretion by human pituitary adenoma cells. Eur J Endocrinol. 2005;152(2):293–300. doi:10.1530/eje.1.01843

54. Iuchi T, Saeki N, Osato K, Yamaura A. Proliferation, Vascular Endothelial Growth Factor Expression and Cavernous Sinus Invasion in Growth Hormone Secreting Pituitary Adenomas. Acta Neurochir (Wien). 2000;142(12):1345–1351. doi:10.1007/s007010070003

55. Galli F, Artico M, Taurone S, et al. Radiolabeling of VEGF165 with 99mTc to evaluate VEGFR expression in tumor angiogenesis. Int J Oncol. 2017;50(6):2171–2179. doi:10.3892/ijo.2017.3989

56. Lee SY, Hong YD, Pyun MS, Felipe PM, Choi SJ. Radiolabeling of monoclonal anti-vascular endothelial growth factor receptor 1 (VEGFR 1) with (177)Lu for potential use in radioimmunotherapy. Appl Radiat Isot Data Instrum Methods Use Agric Ind Med. 2009;67(7-8):1185–1189. doi:10.1016/j.apradiso.2009.02.006

57. Meyer JP, Edwards KJ, Kozlowski P, Backer MV, Backer JM, Lewis JS. Selective Imaging of VEGFR-1 and VEGFR-2 Using 89Zr-Labeled Single-Chain VEGF Mutants. J Nucl Med. 2016;57(11):1811–1816. doi:10.2967/jnumed.116.173237

58. Cai W, Chen K, Mohamedali KA, et al. PET of Vascular Endothelial Growth Factor Receptor Expression. J Nucl Med. 2006;47(12):2048–2056.

59. Barta P, Kamaraj R, Kucharova M, et al. Preparation, In Vitro Affinity, and In Vivo Biodistribution of Receptor-Specific 68Ga-Labeled Peptides Targeting Vascular Endothelial Growth Factor Receptors. Bioconjug Chem. 2022;33(10):1825–1836. doi:10.1021/acs.bioconjchem.2c00272

60. Kniess T. Radiolabeled Small Molecule Inhibitors of VEGFR - Recent Advances. Curr Pharm Des. 18(20):2867–2874. doi:10.2174/138161212800672796

61. Nagengast WB, Vries EG de, Hospers GA, et al. In Vivo VEGF Imaging with Radiolabeled Bevacizumab in a Human Ovarian Tumor Xenograft. J Nucl Med. 2007;48(8):1313–1319. doi:10.2967/jnumed.107.041301

62. Rivadeneira DB, Caino MC, Seo JH, et al. Survivin promotes oxidative phosphorylation, subcellular mitochondrial repositioning, and tumor cell invasion. Sci Signal. 2015;8(389):ra80-ra80. doi:10.1126/scisignal.aab1624

63. Mehrotra S, Languino LR, Raskett CM, Mercurio AM, Dohi T, Altieri DC. IAP regulation of metastasis. Cancer Cell. 2010;17(1):53–64. doi:10.1016/j.ccr.2009.11.021

64. Kong X, Gong S, Su L, et al. Survivin overexpression is potentially associated with pituitary adenoma invasiveness. Oncotarget. 2017;8(62):105637–105647. doi:10.18632/oncotarget.22354

65. Fuchigami T, Mizoguchi T, Ishikawa N, et al. Synthesis and evaluation of a radioiodinated 4,6-diaryl-3-cyano-2-pyridinone derivative as a survivin targeting SPECT probe for tumor imaging. Bioorg Med Chem Lett. 2016;26(3):999–1004. doi:10.1016/j.bmcl.2015.12.046

66. Ishikawa N, Fuchigami T, Mizoguchi T, Yoshida S, Haratake M, Nakayama M. Synthesis and characterization of radioiodinated 3-phenethyl-2-indolinone derivatives for SPECT imaging of survivin in tumors. Bioorg Med Chem. 2018;26(12):3111–3116. doi:10.1016/j.bmc.2018.04.034

