## Supplementary material for "Radiolabeling Molecular Biomarkers of Invasive Pituitary Neuroendocrine Tumors: A Systematic Review": tables

**Tables: See next page.**

| Biomarker | Marker-specific search terms | Shared search term | Papers Reviewed | Papers included/excluded |
|---|---|---|---|---|
| **MMPs** | MMP or metalloproteinase | Invasive or invasiveness<br><br>Pituitary adenoma, pituitary adenomas, pituitary mass, or pituitary tumor | 77 | 5/72 |
| **uPA** | uPA or urokinase-type plasminogen activator | | 2 | 1/1 |
| **MYO5A** | MYO5A, Myosin 5a, Myosin Va (including lowercase variations) | | 1 | 1/0 |
| **VEGF** | VEGF or vascular endothelial growth factor | | 75 | 10/65 |
| **Survivin** | Survivin (including the lowercase variation) | | 11 | 1/10 |

**Table 1**: Search terms for each biomarker reviewed, number of papers retrieved, and papers included and excluded from study.

| Author | Year | Location | Subject | Patient Population | Methods | Results |
|---|---|---|---|---|---|---|
| **Liu HYet al.** | 1996 and 2015/2016 | Multinational | The association between MMP-9 and MMP-2 expression and PitNET invasiveness | 24 Articles<br><br>1,320 patients | Meta-analysis of case-control studies<br><br>Invasiveness was defined by a modified Hardy classification grade of III-IV, Knosp grade III-IV, surgeon-confirmed penetration into adjacent structures, or damage to surrounding tissues<br><br>Identification using either RT-PCR or IHC | MMP-9 expression in invasive tumors was examined by 21 studies, yielding an odds ratio of 5.48 with a 95% confidence interval of 2.61-11.5 (P < 0.00001).<br><br>11 studies: association of MMP-2 expression with invasiveness, reporting an odds ratio of 3.58 and a 95% confidence interval of 1.63-7.87 (P = 0.001).<br><br>An association between the presence of MMP-9 and MMP-2 and the invasiveness of PitNETs has been shown. |
| **Sun Bet al.** | 2020 | UK, Germany, Turkey and China | The association of MMP-9 and PitNET invasiveness | 22 articles<br><br>1,153 Patients: 997 Chinese, 156 in UK, Germany, and Turkey<br><br>Median age 42 years | Meta-analysis<br><br>Positive staining of the nucleus or cytoplasm in pituitary tumor tissue<br><br>Various methods were used by studies to determine invasiveness | 514 of the 631 patients with reported invasive tumors demonstrated positive MMP-9 with an odds ratio of 4.75 and a 95% confidence interval (CI) of 3.53 to 6.39 (p = 0.000).<br><br>Some publication bias was identified in the meta-analysis, as indicated by Begg's and Egger's tests |
| **Guo Het al.** | 2010 and 2012 /2019 | Harrison Int'l Peace Hospital | The association of MMP-9 and invasiveness of pituitary tumors | 108 patients<br><br>61 men and 47 women<br><br>Ranging 25-57 years | Stratification into invasive (n=58) and non-invasive (n=50) groups based on the Hardy-Wilson and Knosp classification systems<br><br>IHC and western blotting for protein-level analysis and RT-PCR for gene-level evaluation | Significant elevation in MMP-9 expression in invasive pituitary tumors compared to non-invasive ones, at both protein and gene levels.<br><br>IHC: MMP-9 positivity rate of 86.21% in invasive tumors, compared to 50% in non-invasive tumors.<br><br>RT-PCR: approximately double the mRNA expression of MMP-9 in invasive tumors relative to non-invasive tumors. Validation by western blotting and serum ELISA, with similar results. |
| **Gong J et al.** | November 2005 to October 2006 /2007 | University of Virginia Medical Center | The association between MMP-9 expression and tumor invasiveness | 73 patients<br><br>37 men, 36 women<br><br>Mean age: 52 years (range 11-79) | MMP-9 expression was analyzed using quantitative RT-PCR, IHC staining, and gelatin zymography, with verification through western blot analysis.<br><br>Invasion is determined by both pathology and gross observation of invasion. | **Table 3 provides a summary of this study's results**. Significant increase in MMP-9 expression in invasive pituitary tumors compared to non-invasive tumors, in functioning and non-functioning tumors. Confirmed through RT-PCR analysis, and gelatin zymography, then validated by IHC staining |
| **Pan L et al.** | 2005 | The people's Hospital of Dongguan | The association between MMP-9 expression and invasiveness | 45 patients<br><br>25 male, 19 female | Biomarker expression was analyzed using IHC staining. Scored based on staining intensity (0–3) by percent of cells involved: 0 = 0%, 1 < 25%, 2 between 26 & 50% 3 > 50%. Invasiveness was determined intraoperatively by the absence of an intact medial wall of the cavernous sinus | The study identified statistically significant associations between pituitary tumor invasiveness and the expression of VEGF (P < 0.001) and MMP-9 (P < 0.001) |

**Table 2**: Descriptions of studies that met the criteria to be analyzed for MMPs 9 and 2. Year column contains information on data collection and research publication. Gong J et al. is described in detail in Table 3.

| Type of PitNET | Technique | Non-Invasive Expression* | Invasive Expression* | *P-Value* |
|---|---|---|---|---|
| **All** | RT-PCR | 0.58 ± 0.09 fold | 2.32 ± 0.38 fold | $P < 0.01$ |
| | Gelatin Zymography | 1.03 ± 0.20 ng/ml | 2.37 ± 0.33 ng/ml | $P < 0.01$ |
| **Functional Tumors** | RT-PCR | 0.52 ± 0.12 fold | 2.55 ± 0.76 fold | $P < 0.01$ |
| | Gelatin Zymography | 1.41 ± 0.19 ng/ml | 3.23 ± 0.65 ng/ml | $P < 0.05$ |
| **Non-Functional Tumors** | RT-PCR | 0.61 ± 0.13 fold | 2.21 ± 0.44 fold | $P < 0.01$ |
| | Gelatin Zymography | 0.77 ± 0.21 ng/ml | 1.94 ± 0.34 ng/ml | $P < 0.01$ |

**Table 3**: The results of Gong J et al, analyzed through various methods, pertaining to all pituitary adenomas, and both functional and non-functional types.

* For RT-PCR, results were evaluated by calculating the fold increase over control, expressed as the ratio of the relative quantities of treated and experimental samples with means calculated via ANOVA. Gelatin zymography results were assessed by comparing activity through densitometric analysis and semi-quantitative measures.

| Author | Year | Location | Subject | Population | Methods | Results |
|---|---|---|---|---|---|---|
| **Sato, M et al.** | 2011-2017 | Keio University School of Medicine | VEGF & VEGFR1 expressions and invasiveness of nonfunctional PitNETs | 27 patients | IHC staining for VEGF and VEGFR1<br><br>Invasiveness determined by CSI* on Gd-enhanced T1 MRI | Statistically significant increases in VEGF and VEGFR1 expression in tumors that invaded the cavernous sinus (*P=0.033* and *P=0.04*, respectively) |
| **Baldys-Waligorska A et al.** | 1997 - 2008 | Jagiellonian University Medical College | VEGF expression levels in a cohort of patients diagnosed with somatotropinomas | 31 patients | Retrospective analysis of clinical, histopathological, and IHC records<br><br>Invasiveness via Knosp grading system & surgeon descriptions. | The analysis did not reveal statistically significant differences in VEGF expression between invasive and non-invasive tumors |
| **He W et al.** | 2017 | 2nd Affiliated Hospital of Nanchang University | The association between VEGF-A expression and the invasiveness of pituitary tumors | 76 patients<br><br>32 males and 44 females | RT-PCR and western blot analyses to<br><br>Invasivness determined by Hardy-Wilson and Knosp grading system. | VEGF-A expression, at both the mRNA and protein levels, was significantly elevated in the invasive pituitary tumor group compared to the non-invasive group |
| **Yilmaz M et al.** | 2003 - 2008 | Kocaeli University Hospital | VEGF expression as a parameter of cavernous sinus invasion | 28 patients | IHC to determine expression.<br><br>Invasiveness defined as Knosp of 4 or 1-3 with: compression of 3+ venous compartments or >45% encasement of ICA. | Statistical analysis using the Chi-Square test revealed a significant positive association between VEGF expression and cavernous sinus invasion ($p = 0.03$) |
| **Sánchez-Ortiga Ret al.** | 1995 - 2008 | Hospital General Universitario de Alicante | VEGF levels of invasive pituitary tumors | 46 pituitary tumor samples.<br><br>39 with extrasellar growth. | RT-PCR<br><br>Invasiveness determined by tumor affecting surrounding structures on presurgical MRI imaging. | Significantly higher normalized copy numbers in tumors with extrasellar growth compared to those without (0.566 vs. 0.200; $p = 0.008$). The study further proposed a normalized copy number threshold of 0.222, which was associated with a 27.5-fold increased risk of extrasellar growth ($p = 0.002$) |
| **Tanase C et al.** | 2013 | Victor Babes National Institute of Pathology | Association between invasiveness & serum VEGF levels in pituitary tumors | 66 samples<br><br>43 invasive & 23 non-invasive | xMAP and ELISA | The study identified significant differences in serum VEGF levels between invasive and non-invasive pituitary tumor |
| **Yarman Set al.** | 2010 | Istanbul University | Expression of VEGF in invasive and non-invasive PAs | 47 GH-secreting PitNETs<br><br>21 invasive & 26 non-invasive | IHC scoring system adding:<br>• Staining Intesity: 0 = negative, 1 = weak, 2 = intermediate, 3 = strong<br>• % cells staining: 0 = 0%, 1 < 25%, 2 < 50%, 3 ≥ 50% | A significant positive association between VEGF positivity and tumor invasiveness ($p = 0.016$) |
| **Borg SA et al.** | 2005 | University of Sheffield Medical School | The relationship between VEGF protein expression and tumor invasiveness | 59 pituitary tumor samples | ELISA to determine expression<br><br>Invasiveness determined by Hardy classification system of 3 & 4 on pre-operative MRI scans | Although invasive tumors demonstrated higher mean VEGF secretion levels compared to non-invasive tumors (2054 ± 1608 pg/mL vs. 608 ± 143 pg/mL), this difference was not statistically significant ($p = 0.867$) |
| **Iuchi T et al.** | 2000 | Chiba Cancer Centre | association between VEGF expression and invasiveness | 25 growth hormone-secreting pituitary tumors | IHC<br><br>Invasiveness determined by Knosp grades 3 and 4 on preoperative imaging. | The study found no statistically significant difference in VEGF expression between invasive and non-invasive growth hormone-secreting pituitary tumors |
| **Pan L et al.** | 2005 | See the last entry in table 2 for more details | | | | |

**Table 4**: Descriptions of studies that met the criteria to be analyzed for VEGF and VEGFR. Year column contains information on data collection and research publication. Pan L et al. is described in detail in Table 2.

* CSI = cavernous sinus invasion
